# Mitochondrial Imaging Detects Early Cardiac Responses to Cancer Immunotherapy

**DOI:** 10.1101/2025.10.13.25337690

**Authors:** Hee-Don Chae, Uttam Shrestha, Millie Das, Minal S. Vasanawala, Deepti Behl, Diwakar Davar, Shyam Srinivas, A. Dimitrios Colevas, John Sunwoo, Niko Del Mar, Michele Pierini, Michele Guillen, Jose Garcia, Negar Omidvari, Nikhil Gupta, Hilda Cabrera, Joseph Blecha, Simon R. Cherry, Henry VanBrocklin, Javid Moslehi, Youngho Seo, Jelena Levi

## Abstract

**One Sentence Summary:** [^18^F]F-AraG PET noninvasively monitors therapy-induced cardiac effects and anti-tumor immunity to guide patient care.

Advances in cancer therapy have improved survival but introduced substantial cardiac risk. Mitochondrial dysfunction underlies cardiotoxicity from conventional therapy, while T cell infiltration drives immunotherapy-related myocarditis. Early detection remains challenging, as current biomarkers and imaging lack sensitivity. Here, we show that [^18^F]F-AraG, a mitochondrial PET tracer that images both activated T cells and cardiomyocytes, may provide an early biomarker of cardiac involvement across different cancer therapies. Healthy cardiac uptake was clearly detectable, consistent across age and sex, and spatially uniform. In patients with cancer, conventional therapy increased cardiac uptake, while immune checkpoint inhibitors induced further increases and regional heterogeneity suggestive of T cell infiltration. Abnormal [^18^F]F-AraG cardiac uptake patterns were observed alongside ECG abnormalities. These findings establish [^18^F]F-AraG PET as a first-in-class imaging tool for simultaneous assessment of early anti-tumor immunity and cancer therapy-related cardiac effects.

## INTRODUCTION

Advances in cancer therapy have substantially improved patient survival, but these gains are often offset by therapy-related toxicities, most notably cardiovascular complications collectively referred to as cardiotoxicity, which have become a leading cause of morbidity and mortality among survivors (*1, 2*). Cardiotoxicity has been associated with chemotherapy, targeted agents, and radiation, with doxorubicin serving as the prototypical example through mechanisms involving oxidative stress, mitochondrial dysfunction, and cardiomyocyte apoptosis (*3*). A unifying feature across these toxicities is mitochondrial injury, which impairs oxidative phosphorylation, redox balance, and biogenesis, ultimately driving apoptosis, fibrosis, and maladaptive remodeling (*4, 5*). Given that mitochondrial dysfunction arises early, before overt structural or functional decline, it represents an attractive diagnostic target for therapy-related cardiotoxicity.

Mitochondria supply >95% of cardiac ATP through fatty acid–driven oxidative phosphorylation and also regulate calcium balance, lipid metabolism, and apoptosis (*6*). Their disruption is therefore both a mechanistic driver of injury and potential window of detection. Mitochondrial biogenesis, i.e. the synthesis of new mitochondria, is tightly coupled to workload and substrate availability, ensuring energy production matches contractile demand (*7*). When dysregulated, this process reduces metabolic flexibility and predisposes the heart to maladaptive remodeling (*8, 9*). Both impaired and compensatory increase in mitochondrial biogenesis may signal early cardiac stress (*10*). Thus, monitoring mitochondrial health, particularly through mitochondrial biogenesis, may help identify therapy-related cardiotoxicity before clinical disease manifests.

Immune checkpoint inhibitors (ICIs) have added a distinct and clinically significant challenge: myocarditis. Although uncommon, ICI myocarditis carries a mortality approaching 50% and is characterized by T cell–mediated myocardial infiltration and cardiomyocyte injury (*11–13*). ICI myocarditis cases are concentrated in melanoma and non–small cell lung cancer (NSCLC), reflecting the primary indications for ICIs (*13*). Most cases occur within weeks of therapy initiation, but early diagnosis is difficult, as current imaging and biomarkers lack sensitivity and specificity for subclinical disease (*14, 15*).

Given these diagnostic limitations, there is an urgent need for a noninvasive, sensitive, and specific tool for the early detection and quantitative assessment of cardiotoxicity. Molecular imaging approaches are under active investigation for this purpose (*16*). Among them, [^18^F]F-AraG, an ^18^F-labeled analog of arabinosylguanine (AraG), possesses distinctive features that make it a particularly compelling candidate for investigation. [^18^F]F-AraG enters cells through nucleoside transporters and is selectively phosphorylated by mitochondrial deoxyguanosine kinase (dGK), allowing its retention in mitochondria and incorporation into mitochondrial DNA, and thereby enabling visualization of cells with high mitochondrial biogenesis via positron emission tomography (PET) (*17–19*). Since mitochondrial biogenesis is critical for T cell activation (*20*), [^18^F]F-AraG PET has been shown to selectively detect activated T cells *in vivo* (*17*). Clinically, [^18^F]F-AraG has been evaluated as a noninvasive imaging tool for systemic immunity in immuno-oncology (*18, 21, 22*). Importantly, its accumulation in mitochondria also enables imaging of cardiomyocytes (*23*).

In this study, we evaluated [^18^F]F-AraG as a potential biomarker for detecting mitochondrial responses in cardiomyocytes and T cell–mediated inflammation during cancer therapy. We first established a stable and clearly discernible baseline of physiological cardiac uptake, which we then used to assess changes in patients with melanoma and in patients with previously treated advanced NSCLC following immunotherapy. Higher overall cardiac uptake was observed in patients with NSCLC who had received prior conventional therapy, which may reflect an adaptive stress response; this pattern was not evident in treatment-naïve stage III melanoma. After ICI therapy, focal increases in cardiac [^18^F]F-AraG uptake, along with elevated global LV uptake, were seen in significant subset of patients in both groups, potentially reflecting immune processes such as T cell infiltration. Taken together, these findings support [^18^F]F-AraG PET as a promising first-in-class approach for simultaneous evaluation of anti-tumor immunity and off-target cardiac effects of cancer therapies, with direct implications for improving patient care.

## RESULTS

### Cardiac [^18^F]F-AraG uptake in cardiac-healthy subjects is stable across different ages and sexes

We conducted PET imaging in 27 healthy individuals to establish baseline physiological levels of cardiac [^18^F]F-AraG uptake. Standardized uptake values (SUV_max_, SUV_mean_) were measured in both the left (LV) and right ventricles (RV). Uptake values were consistent across subjects, as indicated by low coefficients of variation (COV): SUV_max_ – LV: 12.5%, RV: 13.9%; SUV_mean_ – LV: 11.2%, RV: 12.3%, indicating stable tracer distribution in the myocardium (Fig. 1A). One participant with a history of breast cancer therapy demonstrated abnormally elevated cardiac uptake, indicating that prior oncologic treatment may exert persistent cardiac effects despite clinical remission and the absence of overt cardiac disease. Accordingly, this individual was excluded from the healthy control cohort to ensure an unbiased comparison of cardiac uptake patterns with those observed in patients with currently undergoing cancer treatment.

**Fig. 1.**
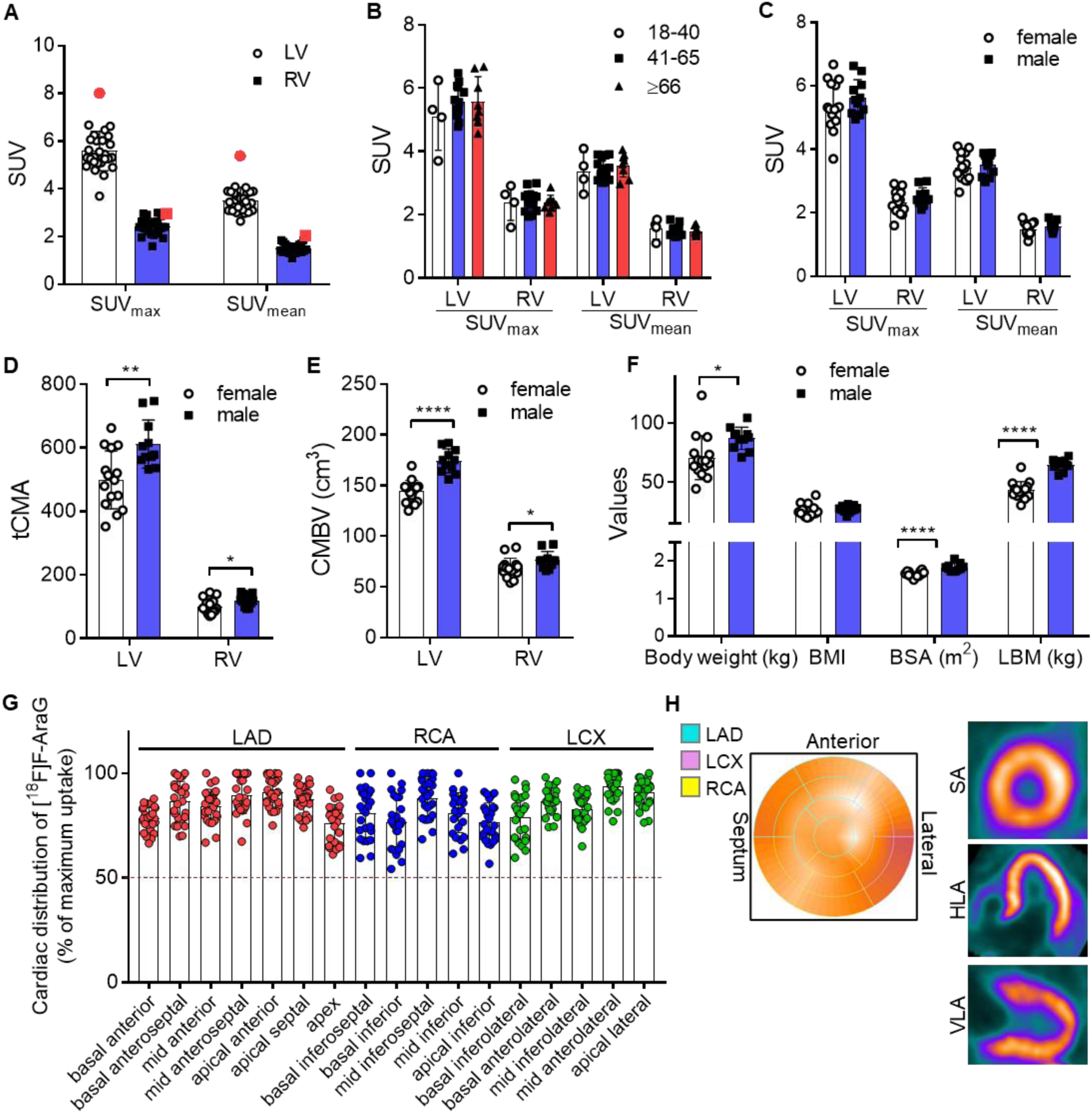
Consistent cardiac [^18^F]F-AraG uptake across age and sex groups. (A) Baseline characterization of cardiac [^18^F]F-AraG uptake in healthy individuals. Healthy participants showed stable LV and RV [^18^F]F-AraG uptake with low COV. One breast cancer survivor with abnormally elevated cardiac uptake (red) was excluded from further analysis. (B–C) Comparison of cardiac [^18^F]F-AraG SUV_mean_ and SUV_max_ between age groups (B) and sexes (C). (D–E) Sex-related differences in tCMA and CMBV. (F) Sex-related differences in body dimensions. (G-H) Regional uniformity of [^18^F]F-AraG uptake within the LV. (G) Myocardial [^18^F]F-AraG uptake was evaluated using the standard 17-segment model and expressed as a percentage of the maximal myocardial uptake per subject. In 26 healthy individuals, uptake was largely homogeneous, with no segments displaying necrotic patterns (defined as uptake <50% of maximum). (H) The representative healthy heart 2-dimensional polar tomogram with reconstructed short axis (SA), vertical long axis (VLA), and horizontal long axis (HLA) slices are displayed together. Data are presented as mean ± SD (age groups: 18–40 (n = 4), 41–65 (n = 14), >65 (n = 8); sex: female (n = 15), male (n = 11)). Statistical analyses were performed by using an unpaired Student t test. *p < 0.05; **p < 0.01; ****p < 0.0001. Each spot represents an individual subject.

Age and sex did not significantly influence cardiac [^18^F]F-AraG SUV_mean_ and SUV_max_ (Fig. 1B and C). To capture the full physiological relevance of [^18^F]F-AraG uptake as a marker of mitochondrial biogenesis, we evaluated two volume-based parameters: total cardiac mitochondrial biogenesis activity (tCMA) and cardiac mitochondrial biogenesis volume (CMBV). tCMA integrates SUV_mean_ with CMBV to quantify overall mitochondrial biogenesis in the myocardium, reflecting the total mitochondrial activity that supports cardiac function and providing a more comprehensive assessment of myocardial health. Notably, we observed significant sex-based differences in these volume-based parameters. Males exhibited higher values than females for both tCMA and CMBV (Fig. 1D and E). This disparity is likely attributable to sex-related differences in body size, such as body weight and body surface area (BSA) (Fig. 1F), which are known to affect cardiac power output (CPO) and heart size (*24–26*). Demographic and [^18^F]F-AraG PET imaging characteristics, including sex comparisons, are summarized in Table 1.

**Table 1.** Demographics and Baseline [^18^F]F-AraG cardiac PET parameters in healthy subjects.

| Parameter | All subjects | Female (n=15) | Male (n=11) | p value |
| --- | --- | --- | --- | --- |
| Median age (range) | 61 (28-76) | 57 (28-76) | 61 (31-73) |  |
| Body weight (kg) | 76.74 ± 17.84 | 69.51 ± 18.86 | 87.25 ± 9.42 | 0.0082 |
| LBM (kg) | 52.26 ± 12.55 | 43.14 ± 7.37 | 64.69 ± 4.82 | 0.00000001 |
| BMI | 25.93 ± 4.50 | 25.28 ± 5.29 | 26.87 ± 2.99 | 0.378 |
| BSA (m<sup>2</sup>) | 1.72 ± 0.12 | 1.65 ± 0.07 | 1.83 ± 0.10 | 0.00002 |
| LV SUV<sub>max</sub> | 5.49 ± 0.68 | 5.39 ± 0.76 | 5.64 ± 0.56 | 0.355 |
| RV SUV<sub>max</sub> | 2.41 ± 0.33 | 2.33 ± 0.36 | 2.51 ± 0.28 | 0.182 |
| LV SUV<sub>mean</sub> | 3.47 ± 0.39 | 3.44 ± 0.43 | 3.51 ± 0.34 | 0.665 |
| RV SUV<sub>mean</sub> | 1.51 ± 0.19 | 1.46 ± 0.20 | 1.57 ± 0.14 | 0.16 |
| LV tCMA | 546.50 ± 100.89 | 499.00 ± 91.57 | 611.26 ± 75.58 | 0.0029 |
| RV tCMA | 109.05 ± 23.77 | 101.06 ± 23.85 | 119.94 ± 19.71 | 0.043 |
| LV CMBV (cm<sup>3</sup>) | 156.96 ± 18.73 | 144.27 ± 10.87 | 174.25 ± 11.88 | 0.000001 |
| RV CMBV (cm<sup>3</sup>) | 71.85 ± 9.78 | 68.60 ± 9.59 | 76.27 ± 8.55 | 0.046 |

We investigated the influence of body size on tCMA and CMBV using a linear regression analysis. LV tCMA was strongly correlated with body weight (r = 0.7261, p < 0.0001) and LBM (r = 0.7363, p < 0.0001), and showed moderate correlations with BSA (r = 0.4856, p = 0.0119) and BMI (r = 0.5756, p = 0.0021) (fig. S1A–D). In contrast, RV tCMA correlated positively only with BSA (r = 0.4673, p = 0.0389) and LBM (r = 0.4924, p = 0.0106) (fig. S1E–H). Similarly, LV CMBV demonstrated strong correlations with body weight (r = 0.7375, p < 0.0001), LBM (r = 0.8971, p < 0.0001), and BSA (r = 0.7399, p < 0.0001), as well as a moderate correlation with BMI (r = 0.4815, p = 0.0127) (fig. S1I). RV CMBV correlated significantly only with BSA (r = 0.4096, p = 0.0378) and LBM (r = 0.4610, p = 0.0178) (fig. S1J–L). Notably, neither tCMA or CMBV differed significantly with age (Nuclear Medicine, Veterans Affairs Palo Alto Health Care System, Palo Alto, CA, USA, 94304 M and N).

### Cardiac-healthy subjects show spatial homogeneity in myocardial [^18^F]F-AraG uptake

The regional homogeneity of tracer distribution within the LV was evaluated using the standard 17-segment model (*27*). Regional analysis showed a uniform distribution, with a coefficient of variation (COV) of 11.11 ± 3.06% among healthy subjects. [^18^F]F-AraG uptake across LV segments averaged 84.16 ± 10.01% relative to the maximum uptake (Fig. 1G). As [^18^F]F-AraG targets mitochondrial DNA (mtDNA) to visualize cardiomyocytes and mitochondria play a fundamental role in cardiomyocyte survival and function (*28*), the absence of [^18^F]F-AraG uptake can serve as a biomarker for cardiomyocyte non-viability (*23*). Regions showing absent or markedly reduced [^18^F]F-AraG uptake, specifically less than 50% of the maximum in the 17-segment analysis, were interpreted as indicative of a necrotic pattern (*29*). Notably, none of the 442 evaluated segments demonstrated necrotic patterns. A two-dimensional polar map confirmed this uniform uptake pattern across the LV, which was also visually consistent in reconstructed short-axis, vertical long-axis, and horizontal long-axis PET views (Fig. 1H). The distribution remained spatially uniform in the breast cancer survivor who exhibited abnormally elevated cardiac [^18^F]F-AraG uptake, suggesting that in this subject cancer therapy altered overall mitochondrial activity without inducing focal regional changes (fig. S2).

### Greater [^18^F]F-AraG uptake in the LV reflects higher mitochondrial biogenesis

The LV pumps blood throughout the entire body under high pressure, whereas the RV delivers blood to the lungs at significantly lower pressure. Due to this disparity in workload, the LV is functionally more powerful than the RV (*30–32*). To meet its higher contractile demands, the heart heavily relies on ATP production, which is primarily generated through mitochondrial oxidative phosphorylation (*6*).

Corresponding to the greater energy requirements of the LV, [^18^F]F-AraG uptake parameters - including SUV_mean_, SUV_max_, and CMV - are approximately twofold higher in the LV compared to the RV, resulting in a fivefold increase in tCMA in the LV (Fig. 2A). Given that AraG is predominantly incorporated into mtDNA (*33*) and [^18^F]F-AraG’s association with mitochondrial biogenesis (*34, 35*), we investigated whether the elevated [^18^F]F-AraG uptake in the LV reflects greater mitochondrial content and biogenesis. To visualize mtDNA, we performed fluorescence in situ hybridization (FISH) on frozen human heart tissue using a human *MT-COX1* sense probe. Stronger mtDNA signals were observed in the LV compared to the RV. Quantitative image analysis confirmed that mtDNA content in the LV was approximately twofold higher than in the RV (Fig. 2B).

**Fig. 2.**
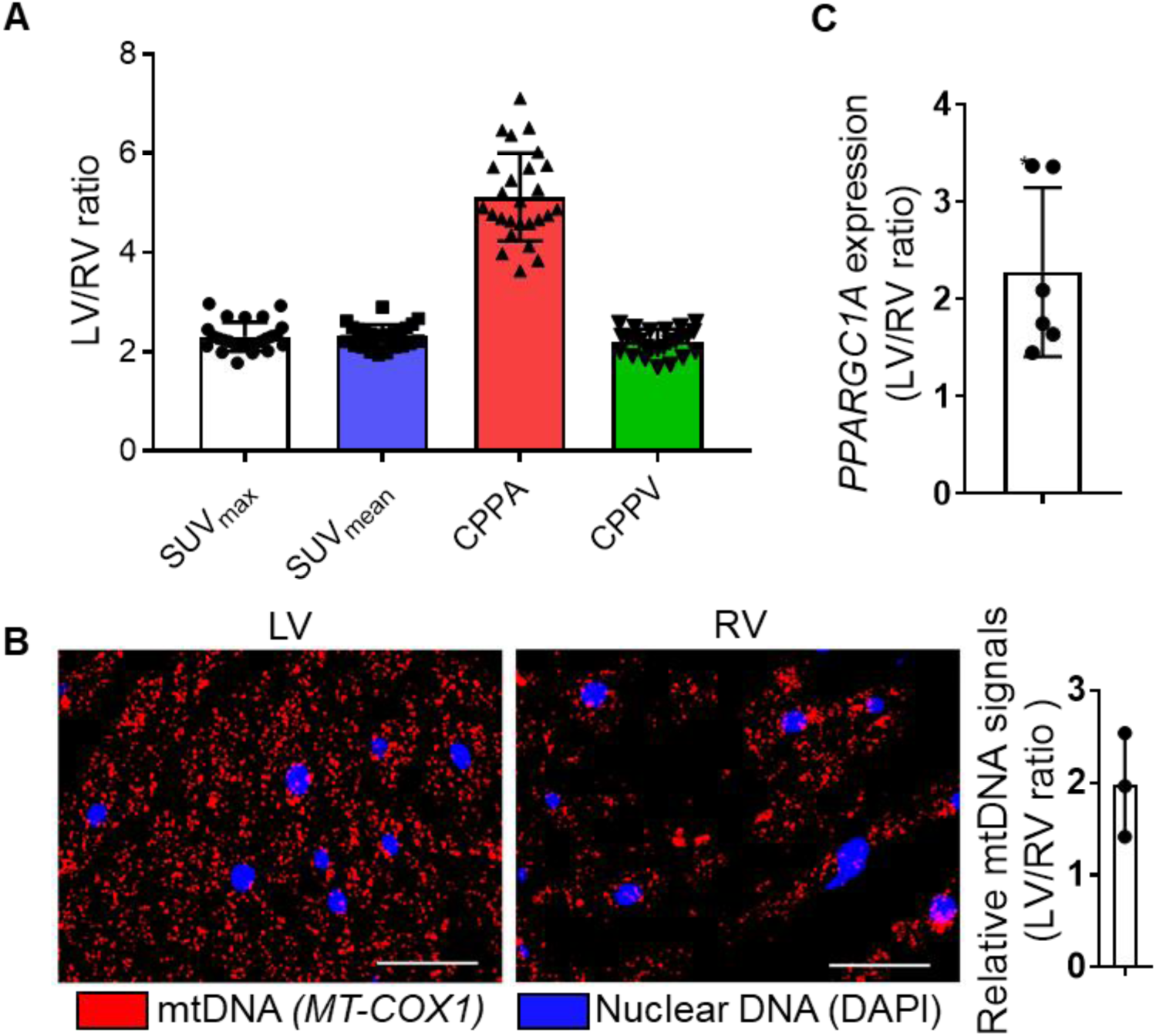
Greater [^18^F]F-AraG uptake in the LV reflects higher mitochondrial biogenesis markers. (A) [^18^F]F-AraG uptake was higher in the LV compared to the RV, reflecting higher work load in LV. Plotted are the LV-to-RV ratios for SUV_max_, SUV_mean_, tCMA, and CMBV values are plotted. (B) Representative mtDNA FISH images showing mtDNA (*MT-COX1*, red) and nuclear DNA (DAPI, blue) in LV and RV tissues (40× magnification; scale bars: 50 µm). FISH image analysis showing higher mtDNA content in LV compared to RV. Corresponding LV/RV mtDNA signal ratios are plotted. Data are presented as mean ± SD (n = 3). (C) *PGC1α*, a key regulator of mitochondrial biogenesis, expression is elevated in LV relative to RV, consistent with increased mitochondrial content. Relative mRNA levels of *PGC1α* were measured by qRT-PCR and normalized to *β-actin* expression. Data are presented as fold-change in LV compared to RV (mean ± SD, n = 6). Statistical analyses were performed by using an unpaired Student t test. *p < 0.05.

To further assess mitochondrial biogenesis, we examined the expression of *PGC-1α* (peroxisome proliferator-activated receptor coactivator-1α), key regulatory protein involved in mitochondrial biogenesis (*36, 37*). Consistent with the increased mtDNA content, *PGC-1α* expression level was significantly higher in the LV than in the RV (Fig. 2C). The observed correlation of [^18^F]F-AraG uptake with mitochondrial content and *PGC-1α* expression highlights its potential as a noninvasive biomarker for assessing mitochondrial biogenesis and homeostasis.

### Increased Myocardial [^18^F]F-AraG Signal Observed After Cancer Therapy

Given that [^18^F]F-AraG uptake in the myocardium primarily occurs in cardiomyocytes rather than T cells in healthy hearts (*23*), the stable cardiac [^18^F]F-AraG uptake provides a reliable baseline for assessing the pathophysiological effects of cancer immunotherapy on cardiac mitochondrial dynamics. Furthermore, the capacity of [^18^F]F-AraG to serve as an in vivo probe for activated T cells (*18, 21, 22*) provides a means to noninvasively monitor myocardial T cell infiltration, a rare yet serious complication of immunotherapy. To address how cancer immunotherapy changes the physiological status of the heart, we retrospectively analyzed [^18^F]F-AraG PET imaging data from two patient groups: treatment-naïve stage III melanoma and previously treated advanced non-small cell lung cancer (NSCLC). [^18^F]F-AraG PET imaging was performed 1-18 days before and 1-4 weeks after the first anti-PD-1 dose. Patient demographics and [^18^F]F-AraG PET parameters are summarized in tables S1 & S2. Quantitative analysis demonstrated that myocardial [^18^F]F-AraG uptake was modestly reduced in therapy-naïve melanoma patients compared with healthy controls (Fig. 3A and table S1). In contrast, patients with advanced NSCLC showed elevated cardiac [^18^F]F-AraG uptake relative to healthy volunteers (Fig. 3A and table S2). To ensure that the observed differences were not attributable to variations in blood pool activity, we compared blood pool uptake between patients with cancer and healthy participants and found no significant differences (fig. S3). While differences between patients with melanoma and NSCLC may reflect differences in cancer type and stage, they most likely result from prior conventional cancer therapy in NSCLC patients, given the known elevated risk of treatment-related cardiovascular adverse effects in patients with NSCLC (*38*). Therapy-induced cardiac stress may trigger an adaptive increase in mitochondrial biogenesis within cardiomyocytes resulting in higher [^18^F]F-AraG signal. Supporting this notion, [^18^F]F-AraG uptake in the LV significantly increased after ICI treatment in patients with melanoma (pre-ICI vs post-ICI, SUV_max_: 4.92 ± 0.53 vs. 5.85 ± 0.81, p < 0.01; SUV_mean_: 2.91 ± 0.31 vs. 3.35 ± 0.44, p < 0.01). Although average group-wise increases in [^18^F]F-AraG uptake were modest in melanoma patients and absent in advanced NSCLC patients, a notable subset of patients showed a significant increase. Specifically, 4 of 7 patients with melanoma (Fig. 3B & fig. S4A) and 4 of 10 patients with NSCLC (Fig. 3C & fig. S4B) showed an over 20% increase in left or right ventricular uptake, surpassing the physiological inter-subject COVs observed in healthy controls. Two patients with NSCLC showed a >20% decrease in RV SUV_max_ following ICI treatment, although their values remained above the average of healthy controls (Fig. 1A & Table 1).

**Fig. 3.**
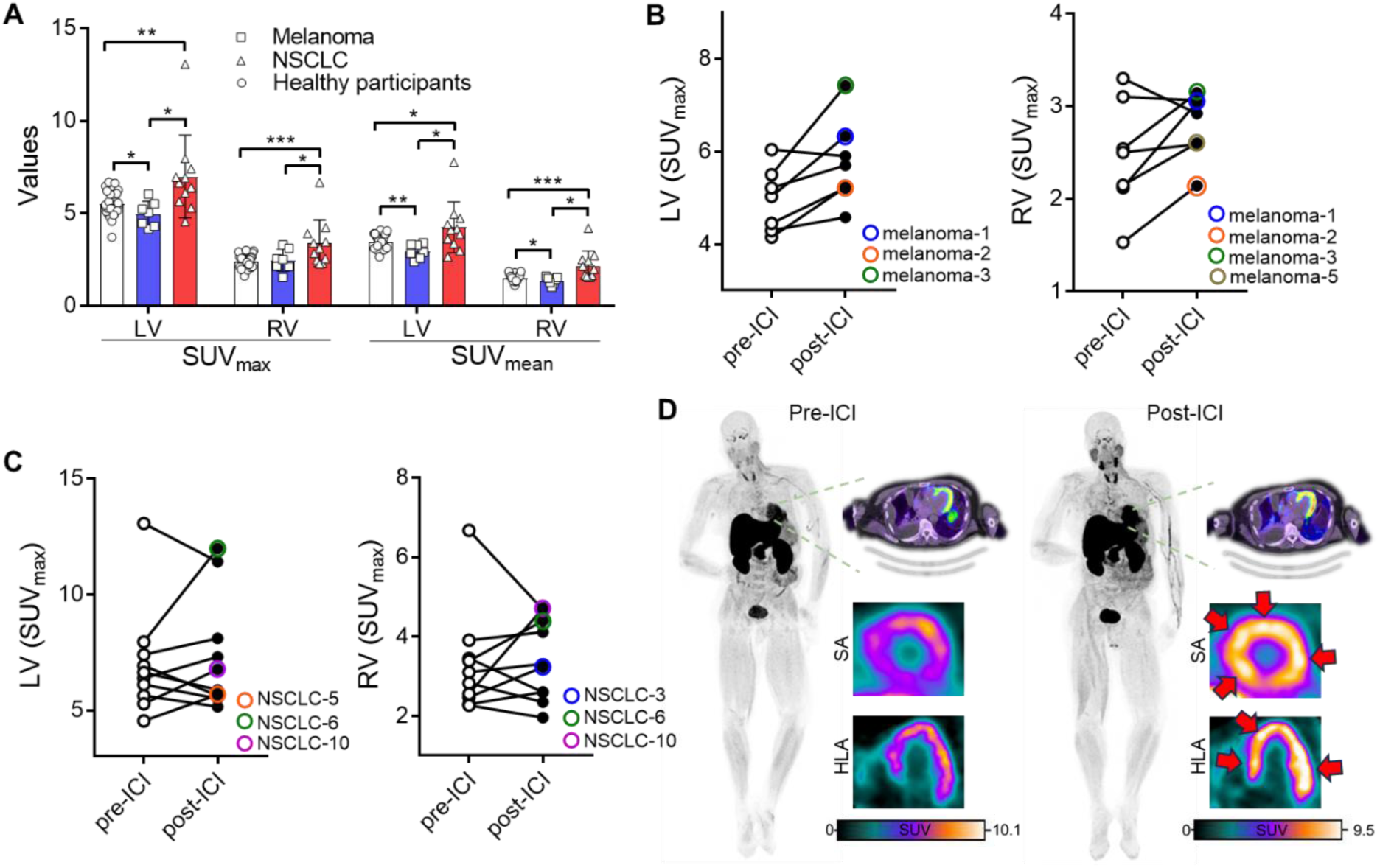
Conventional cancer treatments and immunotherapy induce changes in cardiac [^18^F]F-AraG uptake. (A) Comparison of cardiac SUV_mean_ and SUV_max_ between healthy individuals and patients with melanoma and advanced NSCLC. [^18^F]F-AraG SUV_mean_ and SUV_max_ of were elevated in patients with NSCLC who had received prior therapy, but not in treatment-naïve patients with melanoma. (B) Effect of immunotherapy on cardiac [^18^F]F-AraG uptake in patients with melanoma. Patients with melanoma exhibited a significant increase in cardiac [^18^F]F-AraG SUV_max_ post-ICI. Individuals (4 of 7) who demonstrated more than a 20% increase in either LV or RV [^18^F]F-AraG SUV_max_ are indicated. (C) Effect of immunotherapy on cardiac [^18^F]F-AraG uptake in patients with advanced NSCLC. Patients with >20% increase (4 out of 10) in LV or RV [^18^F]F-AraG SUV_max_ are marked. (D) Representative maximum intensity projection and transaxial PET images from a patient with advanced NSCLC, acquired before and after initiation of Pembrolizumab. Post-ICI treatment, [^18^F]F-AraG uptake increased across multiple tissues, including tumor lesions, lymph nodes, salivary glands, thyroid, and heart, indicating systemic T cell activation. Reconstructed SA and HLA images further demonstrate increased cardiac [^18^F]F-AraG uptake following ICI treatment. Focal [^18^F]F-AraG uptake elevations in the LV after immunotherapy are indicated by red arrows. Groups: Healthy (n = 26); melanoma (n = 7), advanced NSCLC (n = 10). Statistical analyses were performed by using an unpaired (A) or paired (B, C) Student t test. Data are plotted as mean ± SD. *p < 0.05; **p < 0.01; ***p < 0.001.

Interestingly, in some patients, a marked increase in myocardial [^18^F]F-AraG signal following immunotherapy coincided with elevated uptake in tumor lesions, lymph nodes, and several other organs-indicating widespread T cell activation (Fig. 3D). This systemic pattern suggests that the increased cardiac signal may represent not only an adaptive response to ICI treatment but also potential T cell infiltration into the myocardium, a phenomenon occasionally observed with immunotherapy, as has been reported in the context of ICI therapy (*39*). These findings support the potential of [^18^F]F-AraG PET imaging to enable early detection of both off-target cardiac immune-related adverse effects and on-target anti-tumor immune responses associated with cancer immunotherapy.

### Immunotherapy Induces Regional Increases in Cardiac [^18^F]F-AraG Uptake

A homogeneous distribution of [^18^F]F-AraG uptake within the LV is crucial for identifying early regional changes that may indicate cardiovascular risk in cancer patients (*40*). To investigate immunotherapy-associated regional alterations, we analyzed segmental heterogeneity in 7 melanoma and 10 NSCLC patients using the standard 17-segment model (Fig. 4A).

**Fig. 4.**
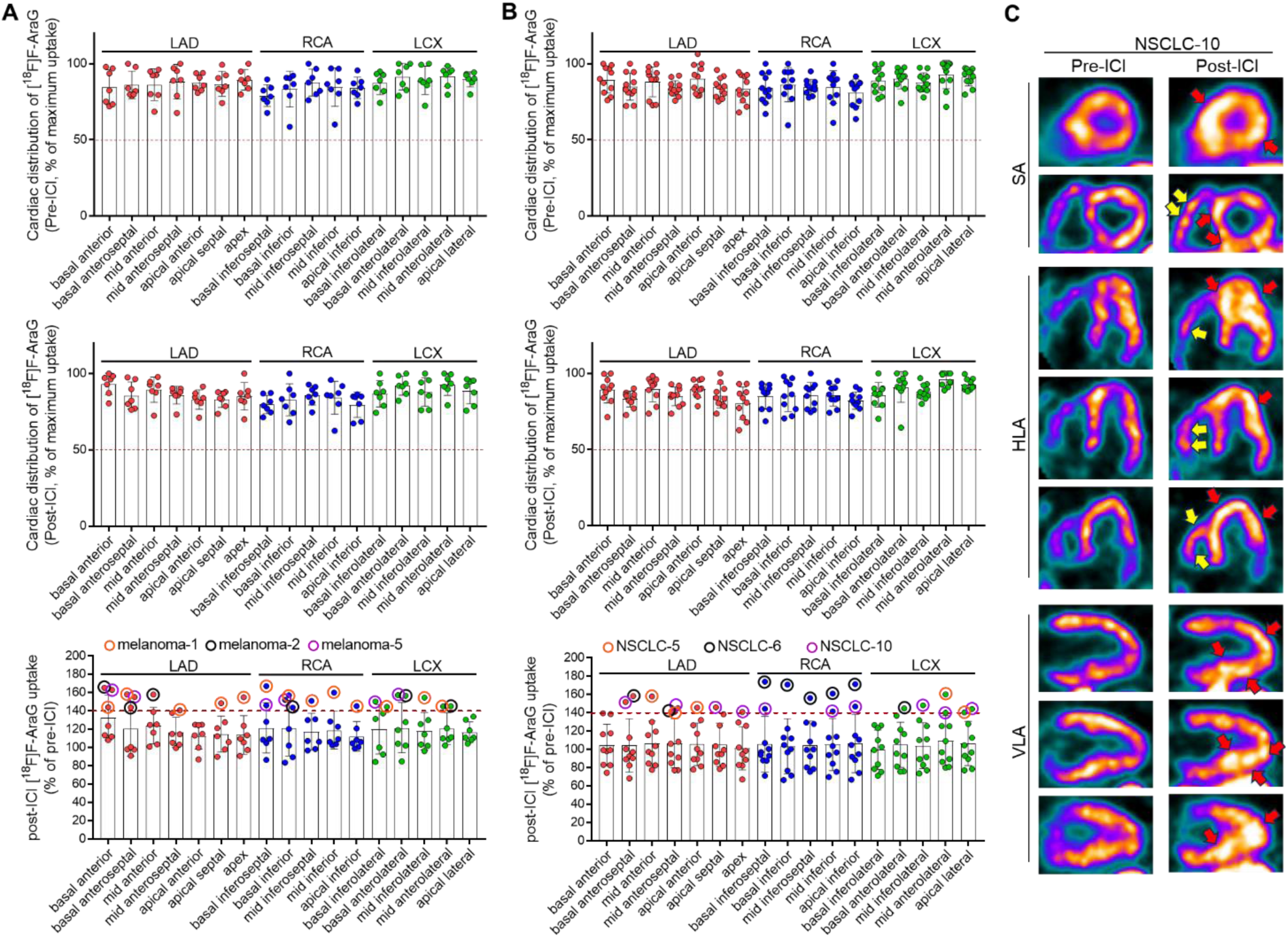
Immunotherapy induced focal increases in cardiac [^18^F]F-AraG uptake. Regional heterogeneity of myocardial [^18^F]F-AraG uptake was evaluated using the 17-segment model in patients with melanoma (A) and advanced NSCLC (B). A subset of patients, 3 out of 7 with melanoma and 3 out of 10 with NSCLC, demonstrated focal increases exceeding 40% in [^18^F]F-AraG uptake in the LV following immunotherapy (see bottom panels). (C) Representative axial [^18^F]F-AraG PET images from patient NSCLC-10 highlight increased focal [^18^F]F-AraG uptake in the RV (yellow arrows) and LV (red arrows) following immunotherapy. Axial images were normalized to their own maximum pixel values for visualizing the relative distribution of the tracer uptake in different myocardial regions.

Across 289 total segments, we observed no necrotic patterns. While overall regional heterogeneity remained relatively stable following ICI treatment (COV (%) was 8.37 ± 2.67 vs. 9.21 ± 2.81 in melanoma patients and 9.29 ± 2.58 vs. 8.60 ± 2.29 in NSCLC patients), a subset of patients (3 of 7 with melanoma and 3 of 10 with NSCLC) showed focal increases in [^18^F]F-AraG uptake exceeding 40% after treatment (Fig. 4A-C). It is notable that most of these patients also had elevated overall LV tracer uptake (Fig. 3 B and C). Because of the retrospective nature of the study, cardiotoxicity-associated biomarkers were not available; nonetheless these results suggest the potential of both [^18^F]F-AraG signal intensity and spatial heterogeneity as early indicators of subclinical immunotherapy-associated cardiac stress.

### Aberrant [^18^F]F-AraG Uptake Pattern Associates with ECG Abnormalities

We evaluated whether myocardial [^18^F]F-AraG uptake could serve as a marker of abnormal cardiac status by comparing it to standard of care cardiac assessment such as electrocardiogram (ECG). In patients with head and neck cancer (H&N) who had available ECG data, heterogeneous myocardial [^18^F]F-AraG uptake was associated with abnormal ECG findings (Fig. 5A). Patient H&N-2 demonstrated a normal ECG and uniform tracer distribution, whereas patient H&N-1 exhibited poor R-wave progression, left axis deviation, and nonspecific T-wave abnormalities in the lateral leads, suggestive of possible anterior infarction. Correspondingly, [^18^F]F-AraG PET imaging of H&N-1 revealed a necrotic pattern in the anterior myocardial segments on 17-segment analysis (Fig. 5B), despite overall LV uptake remaining within the normal range (Fig. 5A). Reconstructed orthogonal PET views further confirmed focal reduction in tracer uptake in the affected areas (Fig. 5C). While this preliminary analysis was limited to data from only two patients, the observations support the potential of [^18^F]F-AraG PET as a noninvasive tool for early detection of cardiotoxicity and assessment of cardiomyocyte viability.

**Fig. 5.**
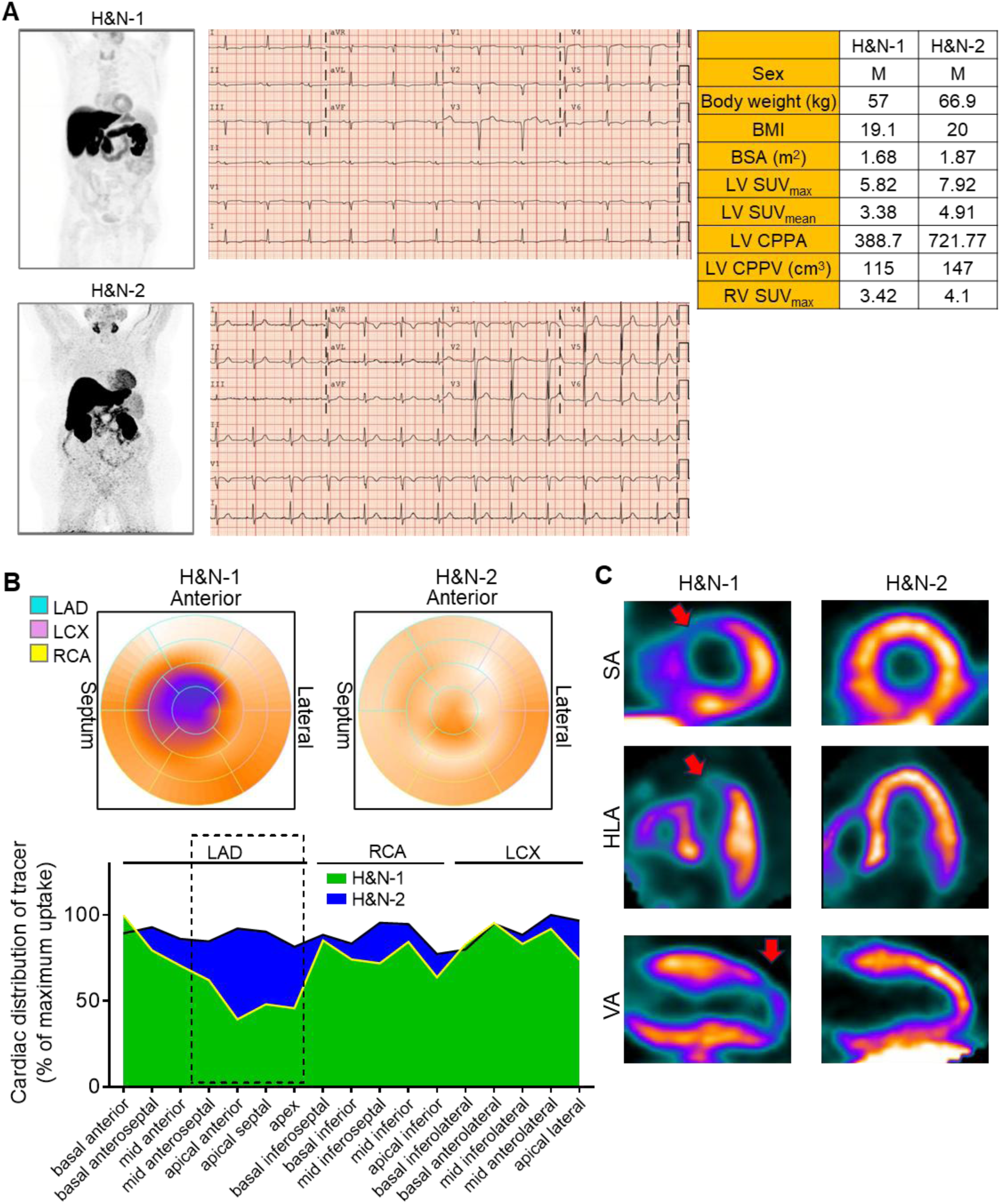
Necrotic pattern of cardiac [^18^F]F-AraG uptake in a patient with head and neck cancer corresponds with ECG abnormalities. (A) ECG tracings and [^18^F]F-AraG maximum intensity projection images from two head and neck cancer patients. Patient H&N-2 exhibited a normal ECG and uniform myocardial [^18^F]F-AraG uptake, whereas patient H&N-1 showed ECG abnormalities accompanied by heterogeneous cardiac uptake. Demographic information and cardiac PET parameters are summarized in the accompanying table. (B) Regional LV [^18^F]F-AraG uptake analysis with the 17-segment model and polar map image of H&N-1 reveal a necrotic pattern characterized by markedly reduced [^18^F]F-AraG uptake in the anterior myocardial segments. (C) Reconstructed SA, VLA, and HLA PET views further confirm the focal reduction of [^18^F]F-AraG uptake in the anterior region of patient H&N-1 (arrows). Reconstructed SA, VLA, and HLA images were normalized to their own maximum pixel values.

## DISCUSSION

Despite significant advances in cancer treatment, therapies like chemotherapy, targeted therapies, and radiation can lead to serious cardiovascular complications. These complications can cause substantial morbidity and mortality in cancer patients, at times exceeding the risks posed by the cancer itself (*1, 2*). Due to the advantage of being noninvasive, cardiac imaging is the most recommended technique for monitoring cardiac function during and after cancer therapy (*41*). Cardiotoxicity from conventional cancer therapies is broadly defined in terms of measurable changes in cardiac function and assessed by serial measurement of left ventricular ejection fraction (LVEF) by echocardiography and multi-gated acquisition. However, a noticeable reduction in LVEF is often a late manifestation of late-stage cardiac injury and may indicate irreversible damage (*14, 15*) and a sizeable proportion of heart failure patients present with preserved LVEF (*42*). Although this modality has advanced and is considered standard, it is not typically sensitive to sub-clinical or early-stage dysfunction (*43*).

Although potentially fatal complications, ICI-related myocarditis remains challenging to predict and diagnose clinically. Symptoms can be nonspecific, ranging from flu-like illness and chest pain to severe heart failure, arrhythmias, or cardiogenic shock. Standard diagnostic markers, such as serum troponin, while almost always elevated, lack specificity as they can also be raised due to skeletal muscle inflammation or other cardiotoxicities (*44*). Patients with ICI myocarditis often present with normal LVEF and a spectrum of clinical HF severity. Cardiac magnetic resonance (CMR) using late gadolinium enhancement allows detection of mechanical changes that occur before left ventricular dysfunction but lacks molecular specificity and sensitivity in ICI-related cardiotoxicity (*14, 15, 43*). While endomyocardial biopsy remains the gold standard for tissue diagnosis, its widespread use is significantly limited by its invasiveness, limited availability in many clinical settings, and a high risk of false negatives (*45*). Given the diagnostic limitations and the rapid, often fatal progression of ICI myocarditis, there is a critical and urgent need for a noninvasive, sensitive, and specific tool to enable early detection and quantitative assessment of this life-threatening condition.

High specificity, resolution, and sensitivity make PET a promising imaging modality for assessment individual biochemical pathways implicated in myocardial pathophysiological molecular pathways such as fibrosis, inflammation, metabolism, mitochondrial function, perfusion, and sympathetic innervation, using radiolabeled probes (*43*). [^18^F]F-FDG PET/CT, widely used for cancer surveillance, has been applied to assess myocardial viability and inflammation in chemotherapy- and ICI-related injury (*16, 46, 47*). However, its interpretation is complicated by the temporal and regional heterogeneity of physiological myocardial glucose uptake. Since the heart’s energy source naturally shifts between fatty acids and glucose depending on the body’s metabolic state, myocardial [^18^F]F-FDG uptake is highly dynamic and influenced by many factors. [^18^F]F-FDG uptake in the heart is strongly influenced by dietary state, fasting conditions, blood glucose levels, myocardial wall stress, underlying disease, and prior therapies (*29, 48*). Although radiolabeled somatostatin analogs such as ^68^Ga-DOTATATE initially showed promise for ICI-myocarditis, their diagnostic performance has proven to be inconsistent (*49*). Given the limited reliability of current methods, efforts are increasingly focused on developing novel radiotracers to advance the diagnostic accuracy and clinical management of cardiotoxicity.

This clinical study on cardiac [^18^F]F-AraG uptake in patients with cancer receiving conventional therapy and immunotherapy validates this emerging mitochondrial imaging biomarker with potential to transform cardio-oncology. This study offers three important insights. First, it establishes a stable, uniform physiological reference for [^18^F]F-AraG uptake in healthy myocardium. Second, it demonstrates the tracer’s ability to noninvasively capture mitochondrial changes in cardiomyocytes in response to conventional therapies. Third, it identifies focal increases in [^18^F]F-AraG uptake, consistent with the hallmark pathological signature of ICI-related myocarditis: patchy, heterogeneous T cell infiltration that is challenging to evaluate with current methods. This capability directly addresses a critical limitation of the gold-standard endomyocardial biopsy, which is prone to sampling error due to the focal nature of immune infiltration. Together, these findings suggest that [^18^F]F-AraG PET may provide a useful approach for early detection and monitoring of cardiac effects induced by cancer therapies.

Our findings are especially relevant given that mitochondrial dysfunction lies at the core of therapy-induced cardiac injury, where disruption of oxidative phosphorylation, elevated reactive oxygen species (ROS), and impaired redox signaling ultimately drive cardiomyocyte apoptosis, fibrosis, and maladaptive remodeling (*4, 5*). Mitochondria possess a robust antioxidant defense system that buffers excessive oxidative stress, thereby protecting cellular components such as DNA, proteins, and lipids from damage that could otherwise lead to cell death and disease (*50*). Sublethal oxidative stress can trigger mitochondrial biogenesis as a protective response. Early molecular events in this process include increased mitochondrial mass and mtDNA content in response to both endogenous and exogenous oxidative stress (*51*). Mitochondrial dysfunction and impaired cardiac energetics are well established as central mechanisms in the development of heart failure. During adaptive hypertrophy, increased mitochondrial biogenesis helps sustain energy production and delays maladaptive decompensation, whereas impaired mitochondrial function accelerates progression toward heart failure (*52*). Elevated cardiac [^18^F]F-AraG uptake in patients with NSCLC receiving conventional therapies (Fig. 3A) suggests that therapy-induced ROS may upregulate mitochondrial biogenesis as an adaptive stress response.

Therapeutic regimens for breast cancer that incorporate anthracyclines, HER2-targeted agents, and radiation are associated with a substantial risk of cardiotoxicity (*4*). Notably, exercise, a potent inducer of mitochondrial biogenesis, has been shown to mitigate LVEF decline in patients treated with doxorubicin and trastuzumab(*5*). In a breast cancer survivor, the pronounced increase in cardiac [^18^F]F-AraG uptake (Fig. 1A) may reflect persistent adaptive responses to previous cancer therapy. In contrast, overwhelming ROS damages mitochondria and other cellular components. This dysfunction amplifies oxidative injury, promotes programmed cardiomyocyte death and fibrosis, and ultimately impairs cardiac function (*4, 5*). Consistent with this, regional analysis in a patient with head and neck cancer revealed a focal decrease in cardiac [^18^F]F-AraG uptake, suggestive of cardiac injury, which was supported by ECG abnormalities. (Fig. 5)

Together, these observations suggest that mitochondrial imaging may offer a valuable approach for understanding and monitoring cardiovascular outcomes in cancer survivors and beyond. By providing a noninvasive, quantitative, and spatially comprehensive assessment of mitochondria and T cell activity in the myocardium, molecular imaging of with cardiac [^18^F]F-AraG PET may enable early detection of cardiotoxicity, guide risk stratification, and monitor interventions aimed at preserving cardiac energetics. At the same time, its ability to capture T cell activation in the tumor microenvironment (*18, 21, 22*) highlights its complementary role in monitoring both cardiotoxicity and anti-tumor immunity.

This preliminary study suggests that [^18^F]F-AraG PET may capture therapy-related changes in cardiac physiology. However, the small sample size, scan heterogeneity, and limited number of patients showing the hypothesized effects restrict the generalizability of these findings. Even so, the consistent uptake patterns observed in healthy participants, together with the therapy-associated changes in patients with cancer, align with established mechanisms of mitochondrial stress and immune-mediated injury. The biological plausibility linking [^18^F]F-AraG to mitochondrial homeostasis and T cell activity further supports the potential relevance of these findings. Standardized multi-center trials incorporating complementary biomarkers such as LVEF, CMR, and troponin will be needed to validate the accuracy and reproducibility of these findings.

## MATERIALS AND METHODS

### Study design

This study aimed to characterize cardiac uptake of [^18^F]F-AraG, a PET tracer that targets mitochondrial DNA to visualize mitochondria-rich cells such as activated T cells and cardiomyocytes, in cancer patients who had undergone treatment, specifically those receiving immune checkpoint inhibitors, with the goal of enabling early detection of therapy-related cardiac adverse events. This objective was achieved by (i) a prospective study to establish baseline physiological cardiac uptake in 26 healthy volunteers, (ii) a retrospective analysis of 7 treatment-naïve patients with stage III melanoma and 10 patients with advanced NSCLC to assess changes in cardiac uptake before and shortly after the first dose of anti-PD-1 therapy, and (iii) a comparison of cardiac [^18^F]F-AraG signals with ECG findings in head and neck cancer patients with available ECG data to evaluate whether abnormal cardiac uptake serves as a marker of pathological cardiac status. To further validate the tracer’s utility for assessing mitochondrial biogenesis and homeostasis, we examined the correlation between cardiac [^18^F]F-AraG uptake, mitochondrial content, and expression of the mitochondrial biogenesis marker *PGC-1α* in the LV and RV.

### Subjects and study protocol

This study was designed prospectively to characterize cardiac uptake of the novel PET tracer [^18^F]F-AraG in healthy individuals and in patients with clinical stage III melanoma (NCT03618641), advanced NSCLC (NCT04726215) or head and neck cancer (NCT03129061). Twenty-seven healthy volunteers (16 females, 11 males; age range 28–76 years) were included in this study. A single outlier in the healthy subject, a breast cancer survivor with abnormally elevated myocardial [^18^F]F-AraG uptake, was excluded from the baseline analyses.

Seventeen cancer patients (melanoma: n = 7; NSCLC: n = 10) were enrolled, with subgroups defined by prior therapy exposure including various immunotherapy treatment. All cancer patients had pre and post therapy (1–4 weeks after receiving their first anti-PD-1 therapy dose) PET/CT acquisition. Four of eight patients with melanoma received intratumoral TLR9 agonist vidutolimod one week prior to treatment with anti-PD1 agent nivolumab (*53*) (table S1). We retrospectively analyzed [^18^F]F-AraG PET images of 2 patients with head and neck cancer scanned in Stanford University.

The study was approved by the Institutional Review Board and Radiation Safety Committee of University of California San Francisco (UCSF), University of California Davis (UC Davis), Stanford University, University of Pittsburgh and other affiliated-participating sites (Sutter Medical Center and VA Palo Alto Healthcare System). All participants provided informed consent in accordance with their participating institution ethics board.

### Image Acquisition and Reconstruction

For the healthy cohort (n = 26), twenty participants were scanned at the UCSF China Basin Imaging Center (NCT02323893) and six at the UC Davis imaging facility (NCT04678440). For cancer patients, PET/CT imaging was performed at Stanford University (2 H&N), University of Pittsburgh (7 melanoma), Sutter Medical Center (5 NSCLC), and VA Palo Alto Healthcare System (5 NSCLC) using standardized protocols with site-specific 3D list-mode image acquisition and vendor-supplied time-of-flight (TOF) reconstruction parameters. All subjects received an intravenous bolus of [^18^F]F-AraG (∼5 mCi). Standard corrections for attenuation, scatter, and radioactive decay were applied to all images. All the reconstructed images were post-processed and smoothed using 3D Gaussian filter.

PET images were acquired approximately one hour (between 50 and 60 minutes) after tracer injection to capture the distribution of [^18^F]F-AraG. A low dose CT scan was included for attenuation correction and anatomical localization with participants in the supine position. At UCSF, healthy volunteers were imaged on a Siemens Biograph64 Vision 600 (slice thickness 4 mm, pixel spacing 1.65 mm) reconstructed with ordered-subset expectation maximization (OSEM; PSF+TOF, 4 iterations, 5 subsets), model-based scatter correction, and a 3D Gaussian filter (FWHM 3.5 mm). At UC Davis, images were acquired on a uEXPLORER total-body PET/CT (2.34 mm isotropic voxel size) (United Imaging Healthcare) reconstructed using a TOF OSEM algorithm with PSF modeling (4 iterations, 10 subsets), Monte-Carlo-based scatter correction, and a 3D Gaussian filter (FWHM 3.5 mm). For the cancer cohort, patients were scanned at additional sites using similar protocols: at University of Pittsburgh and Sutter Medical Center on Siemens Biograph40_mCT 4R systems (slice thickness 4–5 mm, pixel spacing 4 mm) with OSEM (PSF+TOF; 2 iterations, 21 subsets), model-based scatter correction, and a 3D Gaussian filter (FWHM 2.0 mm); at VA Palo Alto Healthcare System on a GE Discovery MI (slice thickness 2.8 mm, pixel spacing 3.64 mm) reconstructed with the CardIQ Function Xpress algorithm, model-based scatter correction, and a 3D Gaussian filter (FWHM 2.9 mm); at Stanford University on a GE Discovery 690 (slice thickness 3.27 mm, pixel spacing 2.08 mm) reconstructed with the OSEM (PSF+TOF; 2 iterations, 21 subsets) algorithm, model-based scatter correction, and a 3D Gaussian filter (FWHM 2.0 mm).

### Image Analysis

PET/CT images were analyzed and quantified using open-source software (Horos, Horos Project; 3D Slicer) with cardiac regional segmentation performed in PMOD (v4.02; PMOD Technologies, Zurich, Switzerland). Standardized uptake values normalized by body weight (SUV_mean_ and SUV_max_) were calculated semi-automatically using a threshold-based segmentation with PET images. Blood pool regions were excluded from LV and RV analysis using PET thresholding to minimize partial-volume effects. LV regional segmentation required reorientation so that the cardiac long axis, from apex to the center of the mitral valve, was orthogonal to the imaging planes, generating standard short-axis, vertical long-axis, and horizontal long-axis views. Myocardial regional segments were defined according to the AHA 17-segment model (*27*). Data were processed using the PMOD cardiac PET analysis tool to reorient LV and RV myocardium along the heart axis by initially supplying a myocardium thickness threshold of 12 mm and manually readjusting slice-by-slice to optimize ROI placement. Uptake patterns and regional heterogeneity were assessed by normalizing each segment to the subject’s maximum myocardial SUV and expressing results as percentages. A necrotic pattern was defined as segmental uptake <50% of the maximum. For visual assessment, myocardial uptake was displayed in polar maps as well as short-axis and long-axis views.

To assess myocardial mitochondrial activity, we calculated: (1) CMBV, defined as volume with uptake ≥50% of SUV_max_; and (2) tCMA, calculated as SUV_mean_ × CMBV. Regional myocardial uptake was mapped to the 17-segment AHA model and expressed as a percentage of each subject’s maximum myocardial SUV.

BSA was calculated using the Du Bois formula (*54*), and LBM using the Janmahasatian equation (55).

### Fluorescence in situ hybridization (FISH) of mtDNA

FISH analysis of mtDNA was performed on fresh frozen human cardiac tissues by VitoVivo Biotech (Rockville, MD). The procedure was conducted manually using the *Hs-MT-COX1*-sense probe in combination with the RNAscope Multiplex Fluorescent Kit v2 (Advanced Cell Diagnostics, Newark, CA), following the manufacturer’s protocol. Human heart tissue samples were obtained from the Duke Human Heart Repository, an IRB-approved tissue bank at Duke University Hospital (*56*). All tissues used in this study were collected from donor hearts deemed unsuitable for transplantation but free of known cardiac pathology.

TSA Vivid Dye 650 was used to label the Hs-MT-COX1 probe, and nuclei were counterstained with DAPI. Images were acquired using a 20× objective and analyzed using Fiji software (*57*). Data were collected from three myocardial regions, with 30 to 97 cells evaluated per region. Quantitative image analysis was used to calculate the number of mtDNA foci per nucleus and to determine the LV-to-RV ratio of mtDNA signals.

### Quantitative reverse transcription polymerase chain reaction (qRT-PCR) analysis of PGC1α Expression in Human Cardiac Tissue

Total RNA was extracted from fresh frozen human cardiac tissue using the QIAGEN RNeasy Plus Universal Mini Kit (Valencia, CA) and reverse-transcribed into cDNA using SuperScript IV VILO Master Mix (Invitrogen, Eugene, OR), following the manufacturer’s instructions. qPCR was performed using PowerTrack SYBR Green Master Mix (Applied Biosystems, Carlsbad, CA) on a QuantStudio 5 384-well system (Applied Biosystems). Primers were selected based on previously published studies (*58, 59*). Primer sequences used were as follows: *PGC1α*, 5′-CCAAAGGATGCGCTCTCGTTCA −3′ and 3′-CGGTGTCTGTAGTGGCTTGACT −5′; *ACTB*, 5′-GGACTTCGAGCAAGAGATGG −3′ and 5′-AGCACTGTGTTGGCGTACAG −3′. PGC1α expression levels were normalized to β-actin, and relative expression was calculated using the 2^−ΔΔCT^ method (*60*) and the left-to-right ventricular (LV/RV) expression ratio of *PGC1α* was determined.

### Statistical analysis

All data are reported as the mean ± standard deviation (SD). Paired and unpaired Student’s t-tests were used where appropriate. A p value less than .05 was considered to indicate a significant difference.

Statistical analyses and Pearson correlation analyses were performed using GraphPad Prism (Version 7.05, GraphPad Software, La Jolla, CA).

## List of Supplementary Materials

Fig S1 to Four Table S1 to Two

## Data Availability

All data associated with this study are present in the paper or the Supplementary Materials.

## Acknowledgments

We thank the Duke Human Heart Repository, led by Dr. Carmelo Milano and Dr. Dawn Bowles, for generously providing human cardiac tissue.

## Author Contributions

J.D. and Y.S. acquired funding and supervised all aspects of the study. J.D., Y.S., U.S., and H.-D.C. conceived the experiments and analyzed the data. M.D., M.S.V., D.B., D.D., S.S., A.D.C., J.S., N.D.M., M.P., M.G., J.G., N.O., H.C., and S.R.C. recruited and clinically characterized study participants and acquired clinical data and PET images. J.B., S.R.C., H.V., and J.M. provided expert advice and guidance on data interpretation. N.G. interpreted the ECG data. U.S. analyzed the PET data.

H.-D.C. interpreted the data and designed the figures. H.-D.C. and U.S. wrote the first draft of the manuscript. All authors critically revised the manuscript and approved the final version.

## Data and materials availability

All data associated with this study are present in the paper or the Supplementary Materials.

