## Supplementary material for "Mitochondrial Imaging Detects Early Cardiac Responses to Cancer Immunotherapy": fig. S1, fig. S2, fig. S3, fig. S4, Table S1, Table S2

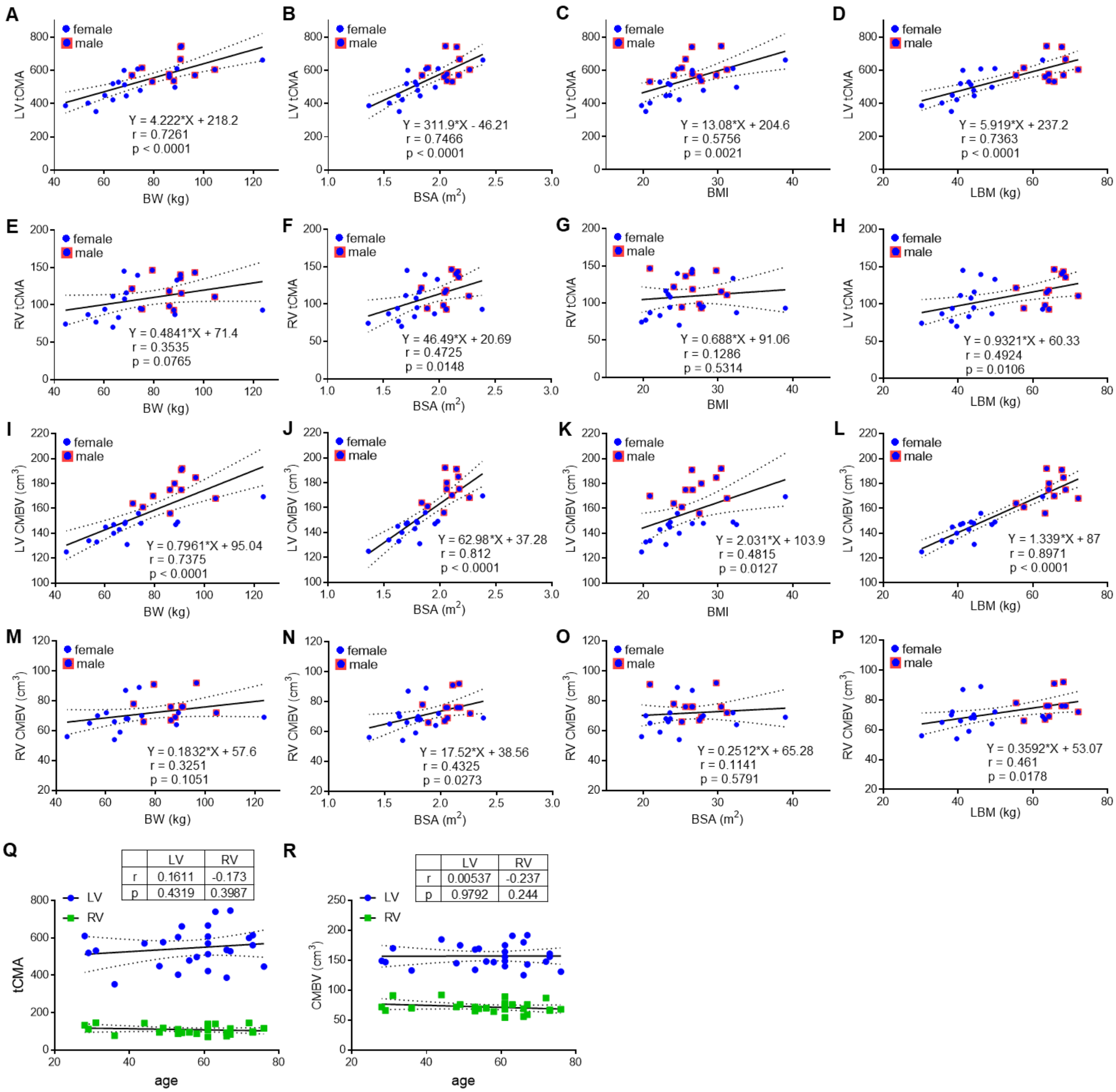


**Fig. S1. Effect of body size parameters and age on total cardiac mitochondria biogenesis activity (tCMA) and cardiac mitochondria biogenesis volume (CMBV).** Scatter plots depict linear correlations between total cardiac mitochondrial activity (tCMA) or mitochondrial biogenesis volume (CMBV) parameters. tCMA in the LV shows positive correlations with BW **(A)**, BSA **(B)**, BMI **(C)**, and LBM **(D)**. Right ventricular tCMA correlates with BSA **(F)** and LBM **(H)**, but not with BW **(E)** or BMI **(G)**. LV CMBV positively correlates with BW **(I)**, BSA **(J)**, BMI **(K)**, and LBM **(L)**, while RV CMBV shows a correlation only with BSA **(N)** and LBM **(P)**. Neither BW nor BMI is associated with RV tCMA or RV CMBV. Age did not influence both tCMA and CMBV **(Q, R)**. female (n = 15), male (n = 11).

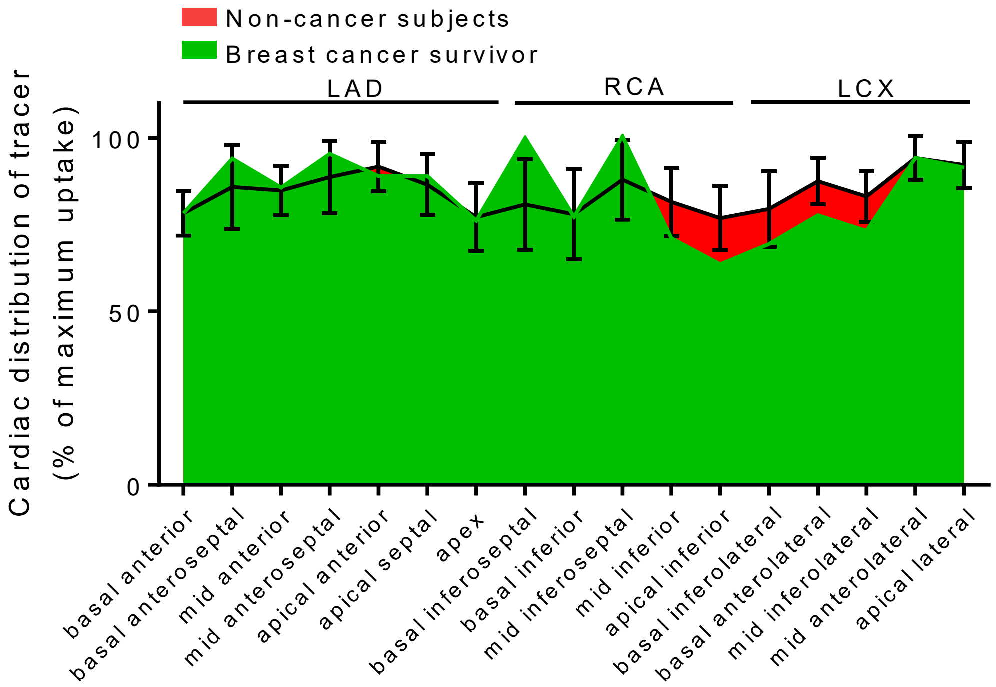


**Fig. S2. Uniform cardiac [$^{18}$F]F-AraG uptake in a breast cancer survivor.** Although cancer therapy may have induced physiological changes leading to increased overall cardiac [$^{18}$F]F-AraG uptake, spatial distribution remained homogeneous, as demonstrated by analysis using the standard 17-segment cardiac model.

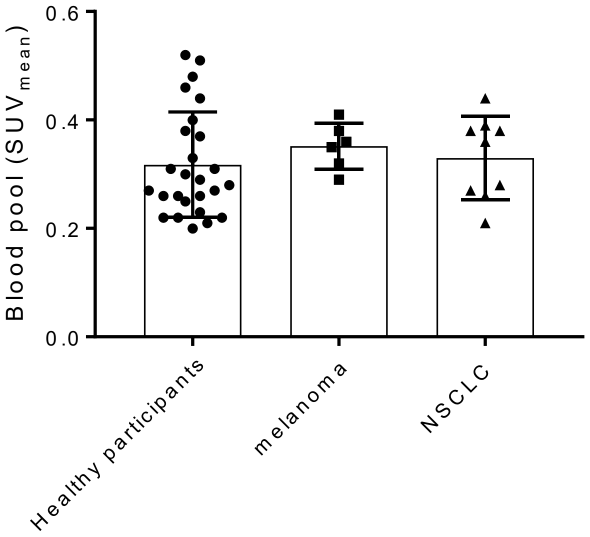


**Fig. S3. Comparable blood pool [$^{18}$F]F-AraG uptake in healthy and cancer patients.** The blood pool $SUV_{mean}$ was measured using the volume of interest drawn manually on the center of the ascending aorta. Healthy (n = 26); melanoma (n = 6), advanced NSCLC (n = 8). Blood pool $SUV_{mean}$ was excluded in four patients: one with melanoma (due to high noise near the aorta) and two with NSCLC (due to spillover from adjacent tumors).

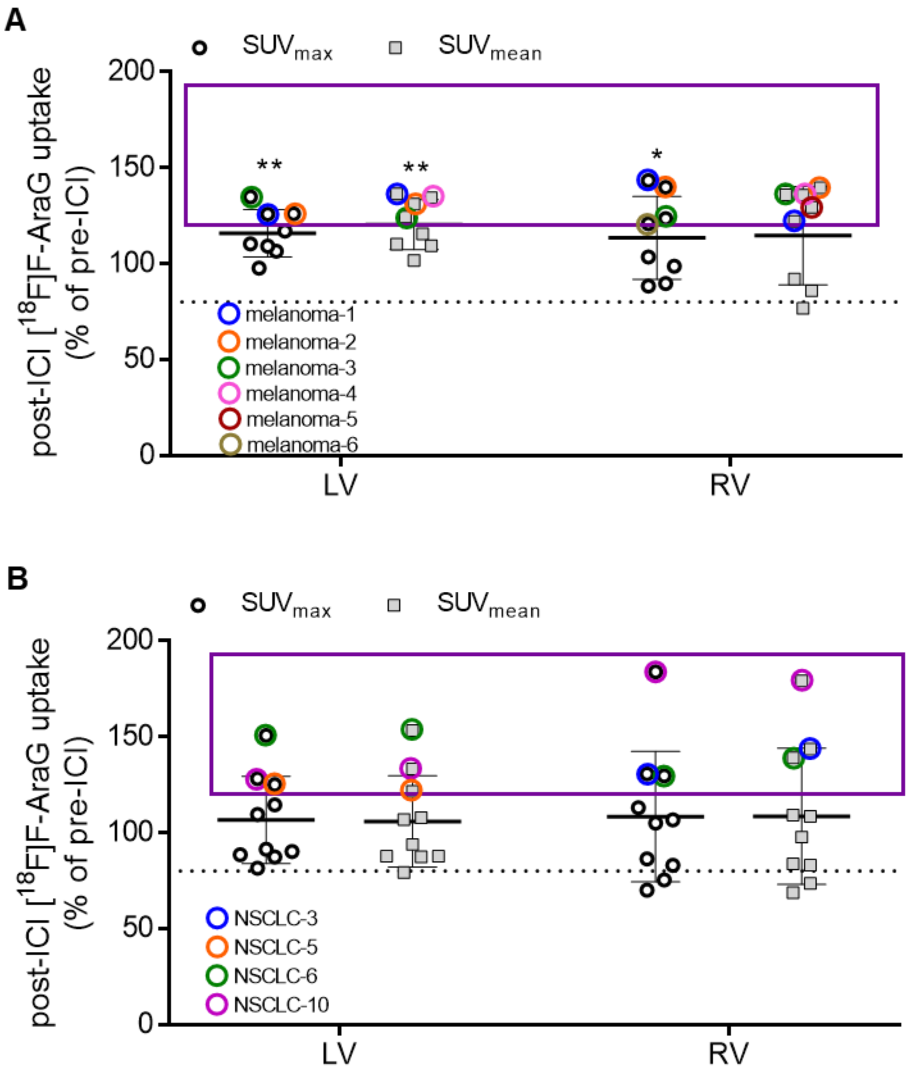


**Fig. S4. Elevated cardiac [18F]F-AraG uptake following ICI treatment. (A)** Effect of immunotherapy on cardiac [18F]F-AraG uptake in patients with melanoma. Patients with melanoma exhibited a significant increase in cardiac [18F]F-AraG uptake post-ICI. Individuals who demonstrated more than a 20% increase in either LV or RV uptake are indicated. **(B)** Effect of immunotherapy on cardiac [18F]F-AraG uptake in patients with advanced NSCLC. Patients with >20% increase in LV or RV uptake are marked. Groups: Healthy (n = 26); melanoma (n = 7), advanced NSCLC (n = 10). Statistical analyses were performed by using a paired Student t test. Data are plotted as mean ± SD. $^{**}p < 0.01$.

**Table S1. Demographics and cardiac [$^{18}$F]F-AraG PET metrics before and after immune checkpoint inhibitor therapy in melanoma subjects.**

| | melanoma-1 | | melanoma-2 | | melanoma-3 | | melanoma-4 | | melanoma-5 | | melanoma-6 | | melanoma-7 | |
|---|---|---|---|---|---|---|---|---|---|---|---|---|---|---|
| Age | ≥66 | | 41-65 | | ≥66 | | ≥66 | | ≥66 | | ≥66 | | ≥66 | |
| Sex | M | | F | | M | | M | | M | | F | | M | |
| Body weight (kg) | 88.9 | | 56.7 | | 118.8 | | 133.8 | | 80.49 | | 85.02 | | 80.74 | |
| BMI | 30.8 | | 22.2 | | 38.8 | | 40.0 | | 26.3 | | 32.4 | | 26.4 | |
| BSA (m$^2$) | 1.98 | | 1.59 | | 2.36 | | 2.51 | | 1.96 | | 1.90 | | 1.97 | |
| prior therapy | N | | N | | N | | N | | N | | N | | N | |
| Vidutolimod | Y | | Y | | N | | N | | N | | N | | Y | |
| ICI therapy | Nivolumab | | Nivolumab | | Nivolumab | | Nivolumab | | Nivolumab | | Nivolumab | | Nivolumab | |
| ICI to post PET | 16d | | 15d | | 28d | | 24d | | 15d | | 11d | | 15d | |
| | pre ICI | post ICI | pre ICI | post ICI | pre ICI | post ICI | pre ICI | post ICI | pre ICI | post ICI | pre ICI | post ICI | pre ICI | post ICI |
| LV $SUV_{max}$ | 5.03 | 6.33 | 4.15 | 5.23 | 5.50 | 7.42 | 6.04 | 5.90 | 4.60 | 4.50 | 5.22 | 5.70 | 4.45 | 5.21 |
| RV $SUV_{max}$ | 2.12 | 3.04 | 1.53 | 2.14 | 2.54 | 3.14 | 3.30 | 2.92 | 2.15 | 2.60 | 2.50 | 2.59 | 3.10 | 3.06 |
| LV $SUV_{mean}$ | 2.91 | 3.97 | 2.38 | 3.12 | 3.40 | 4.23 | 3.28 | 3.34 | 2.85 | 2.98 | 3.32 | 3.63 | 2.83 | 3.12 |
| RV $SUV_{mean}$ | 1.18 | 1.44 | 1.01 | 1.41 | 1.42 | 1.93 | 1.53 | 1.98 | 1.59 | 1.22 | 1.26 | 1.16 | 1.49 | 1.28 |
| LV tCMA | 637.29 | 960.74 | 323.68 | 486.72 | 656.20 | 854.46 | 695.36 | 631.26 | 592.80 | 634.74 | 564.40 | 613.47 | 625.43 | 670.80 |
| RV tCMA | 69.62 | 87.84 | 56.56 | 83.19 | 97.98 | 140.89 | 113.22 | 142.56 | 108.12 | 85.40 | 76.86 | 77.72 | 87.91 | 75.52 |
| LV CMBV (cm$^3$) | 219 | 242 | 136 | 156 | 193 | 202 | 212 | 189 | 208 | 213 | 170 | 169 | 221 | 215 |
| RV CMBV (cm$^3$) | 59 | 61 | 56 | 59 | 69 | 73 | 74 | 72 | 68 | 70 | 61 | 67 | 59 | 59 |

**Table S2. Demographics and cardiac [$^{18}$F]F-AraG uptake parameters before and after immune checkpoint inhibitor therapy in patients with NSCLC.**

| | NSCLC-1 | | NSCLC-2 | | NSCLC-3 | | NSCLC-4 | | NSCLC-5 | | NSCLC-6 | | NSCLC-7 | | NSCLC-8 | | NSCLC-9 | | NSCLC-10 | |
|---|---|---|---|---|---|---|---|---|---|---|---|---|---|---|---|---|---|---|---|---|
| PET Center[a] | PAVA | | PAVA | | PAVA | | PAVA | | PAVA | | Sutter | | Sutter | | Sutter | | Sutter | | Sutter | |
| Age | ≥66 | | ≥66 | | ≥66 | | ≥66 | | ≥66 | | 41-65 | | 41-65 | | ≥66 | | ≥66 | | ≥66 | |
| Sex | M | | M | | M | | M | | M | | M | | F | | F | | F | | M | |
| Body weight (kg) | 69.7 | | 93.5 | | 92.5 | | 45.7 | | 84.4 | | 61.7 | | 61.7 | | 86.63 | | 68.3 | | 86.63 | |
| BMI | 25.9 | | 30.9 | | 29.5 | | 15.8 | | 26.9 | | 26.6 | | 25.7 | | 34.9 | | 27.5 | | 25.2 | |
| BSA (m$^2$) | 1.76 | | 2.08 | | 2.10 | | 1.51 | | 2.02 | | 2.21 | | 1.60 | | 1.87 | | 1.69 | | 2.11 | |
| prior therapy | Y | | Y | | Y | | Y | | Y | | Y | | Y | | Y | | Y | | Y | |
| ICI therapy | Pembrolizumab | | Pembrolizumab | | Pembrolizumab | | Atezolizumab | | Pembrolizumab | | Pembrolizumab | | Pembrolizumab | | Pembrolizumab | | Pembrolizumab | | Pembrolizumab | |
| ICI to post PET | 6d | | 18d | | 9d | | 6d | | 6d | | 7d | | 10d | | 16d | | 8d | | 11d | |
| | pre ICI | post ICI | pre ICI | post ICI | pre ICI | post ICI | pre ICI | post ICI | pre ICI | post ICI | pre ICI | post ICI | pre ICI | post ICI | pre ICI | post ICI | pre ICI | post ICI | pre ICI | post ICI |
| LV $SUV_{max}$ | 7.40 | 8.11 | 6.92 | 5.64 | 6.38 | 7.31 | 6.11 | 5.51 | 4.54 | 5.68 | 7.95 | 11.98 | 6.64 | 5.88 | 13.06 | 11.41 | 5.63 | 5.15 | 5.28 | 6.77 |
| RV $SUV_{max}$ | 3.91 | 4.11 | 3.45 | 2.60 | 2.47 | 3.23 | 3.10 | 3.31 | 2.30 | 2.60 | 3.39 | 4.40 | 2.27 | 1.96 | 6.67 | 4.68 | 2.83 | 2.35 | 2.56 | 3.52 |
| LV $SUV_{mean}$ | 4.96 | 5.30 | 4.73 | 3.75 | 4.15 | 4.48 | 4.11 | 3.59 | 2.97 | 3.61 | 4.41 | 6.76 | 3.62 | 3.40 | 7.76 | 6.81 | 3.35 | 2.94 | 2.64 | 3.52 |
| RV $SUV_{mean}$ | 2.49 | 2.72 | 2.43 | 1.79 | 1.54 | 2.21 | 2.15 | 2.10 | 1.63 | 1.77 | 1.97 | 2.74 | 1.53 | 1.27 | 4.19 | 2.88 | 1.66 | 1.39 | 1.59 | 2.85 |
| LV tCMA | 848.16 | 911.60 | 827.75 | 637.50 | 871.50 | 887.04 | 530.19 | 416.44 | 531.63 | 678.68 | 776.16 | 1216.80 | 485.08 | 482.80 | 1125.20 | 919.35 | 576.20 | 552.72 | 366.96 | 513.92 |
| RV tCMA | 174.30 | 198.56 | 172.53 | 127.09 | 147.84 | 143.65 | 118.25 | 111.30 | 101.06 | 116.82 | 116.23 | 147.96 | 82.62 | 71.12 | 196.93 | 132.48 | 84.66 | 73.67 | 84.27 | 153.90 |
| LV CMBV (cm$^3$) | 171 | 172 | 175 | 170 | 210 | 198 | 129 | 116 | 179 | 188 | 176 | 180 | 134 | 142 | 145 | 135 | 172 | 188 | 139 | 146 |
| RV CMBV (cm$^3$) | 70 | 73 | 71 | 71 | 96 | 65 | 55 | 53 | 62 | 66 | 59 | 54 | 54 | 56 | 47 | 46 | 51 | 53 | 53 | 54 |

a: VA Palo Alto Healthcare System (PAVA), Sutter Medical Center (Sutter)
